# LITERATURE REVIEW THE RELATIONSHIP BETWEEN NURSES RESILIENCE AND SOCIAL SUPPORT : PHILOSOPHY OF NURSING

**DOI:** 10.1101/2024.11.29.24318229

**Authors:** Naya Ernawati, Moses Glorino Rumambo Pandin, Ah Yusuf

## Abstract

**Introduction:** The purpose of this study is to examine the philosophy of social support from the perspective of the resilience.

**Methods:** This study used a literature review using the PICOS framework. There are 11 articles in it, articleswere searched from 4 databases namely Sciencedirect, Scopus, PubMed, CINAHL. The keywords used in the literature search were resilience AND social support, AND nurses AND hospital. The search was limited to 2019-2024 publications, full articles, not review articles. The strategy used to search for articlesusing the PICOS framework. Then selected using the PRISMA diagram andobtained eleven articles. Quality assessment of eleven articles using the JBI CriticalAppraisal tool.

**Results:** Ten articles describing the relationship between social support and resilience. Social Support can be used improve resilience in nurses at hospital.

**Conclusion:** Social support is related to psychological resilience. Building resilience and social support can help reduce nurses’ vulnerability. Policymakers must develop and implement appropriate strategies to increase nurses’ social support and resilience to improve the quality of nursing care.

## INTRODUCTION

Improvements to modern health services requires health workers to be tough to face difficult situations (WHO, 2021a). Some people are more ‘resilient’ than others, so they tend to be able to cope with difficult times better, thus remaining in the workplace. However, the wider literatureconcerns staff resilience It seems more than just ‘running out of steam’; it involves positive adaptation and development of personal resources (Cooper et al., 2021). Poor workplace conditions can affect professional resilience and building resilience among nurses have been considered to support staff retention and reduce their turnover (Ahmed et al., 2022;. Zhang et al. (2020) reported a significant negative correlation between resilience and burnout in healthcare circles.

Various other factors can contribute to people’s burnout and resilience when exposed to high job demands and stressful work environments among those are organisational and social support. Employees’ perception of organisational support is based on how much their employers consider their needs and value their work (Eisenberger et al., 1986). High levels of perceived organisational support among healthcare workers have been positively correlated with reducing work- related stress and increasing the quality of patient care, staff performance, job satisfaction and commitment (de los Santos & Labrague, 2021; Ture & Akkoc, 2021). Low levels of perceived organisational support are associated with work absenteeism and intent to turnover (Wang & Wang, 2020). Social support is equally important, and it is related to the quality and function of social relationships that one receives from other people. There are different sources of social support such as family and friends. Social support helps individuals develop close social bonds with other people, provides a sense of belonging and supports people feeling that their life is meaningful. Having adequate social support help people cope with adversities in their life, enhance people’s health, well-being, resilience and reduce the risk of burnout (Zhang et al., 2020).

Nurses had a low degree of psychological resilience in to begin with. Nurses self-reported that they would have more confidence in their capacity to relieve stress if felt supported by family and friends (Chigwedere et al., 2021; Huang et al., 2021). Almazan et al.’s study of older adults who had survived disasters found that social support could help increase the psychological resilience of older adults in the face of disasters (Almazan et al., 2019).Because social support can improve psychological resilience, and psychological resilience is a positive factoron quality of life, psychological resilience may play a mediating role between social support and quality of life.

## MATERIALS AND METHODS

The literature for the originality of this study used articles in English from 4 databases: Sciencedirect, Scopus, PubMed, CINAHL. The keywords used in the literature search were: the literature search were resilience AND social support, AND nurses AND hospital. The strategy used to searchfor articles usingthe PICOS framework consists of:

**Figure 1.**
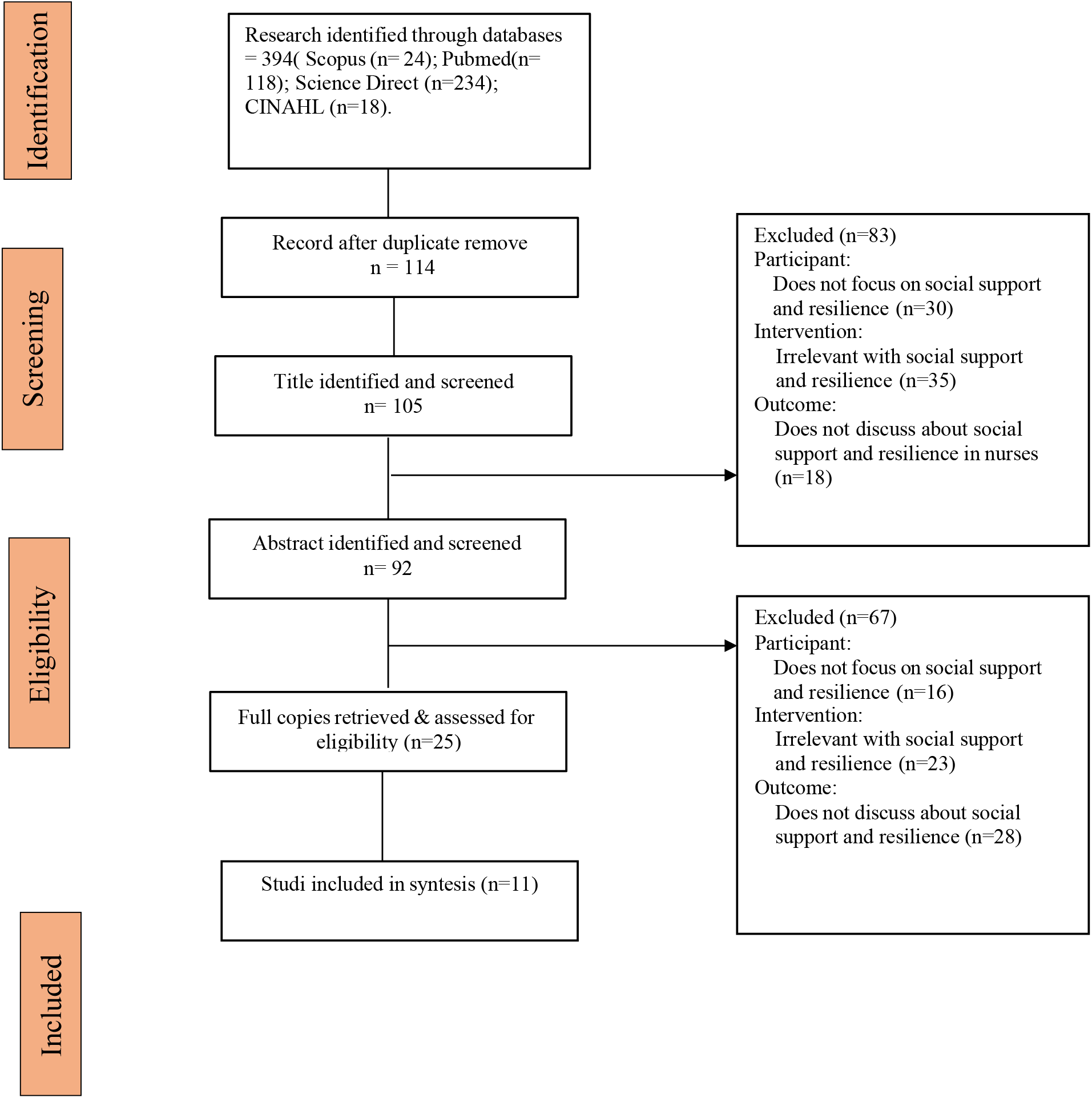
Literature Search Flow Diagram based on PRISMA.

**Table 1.**
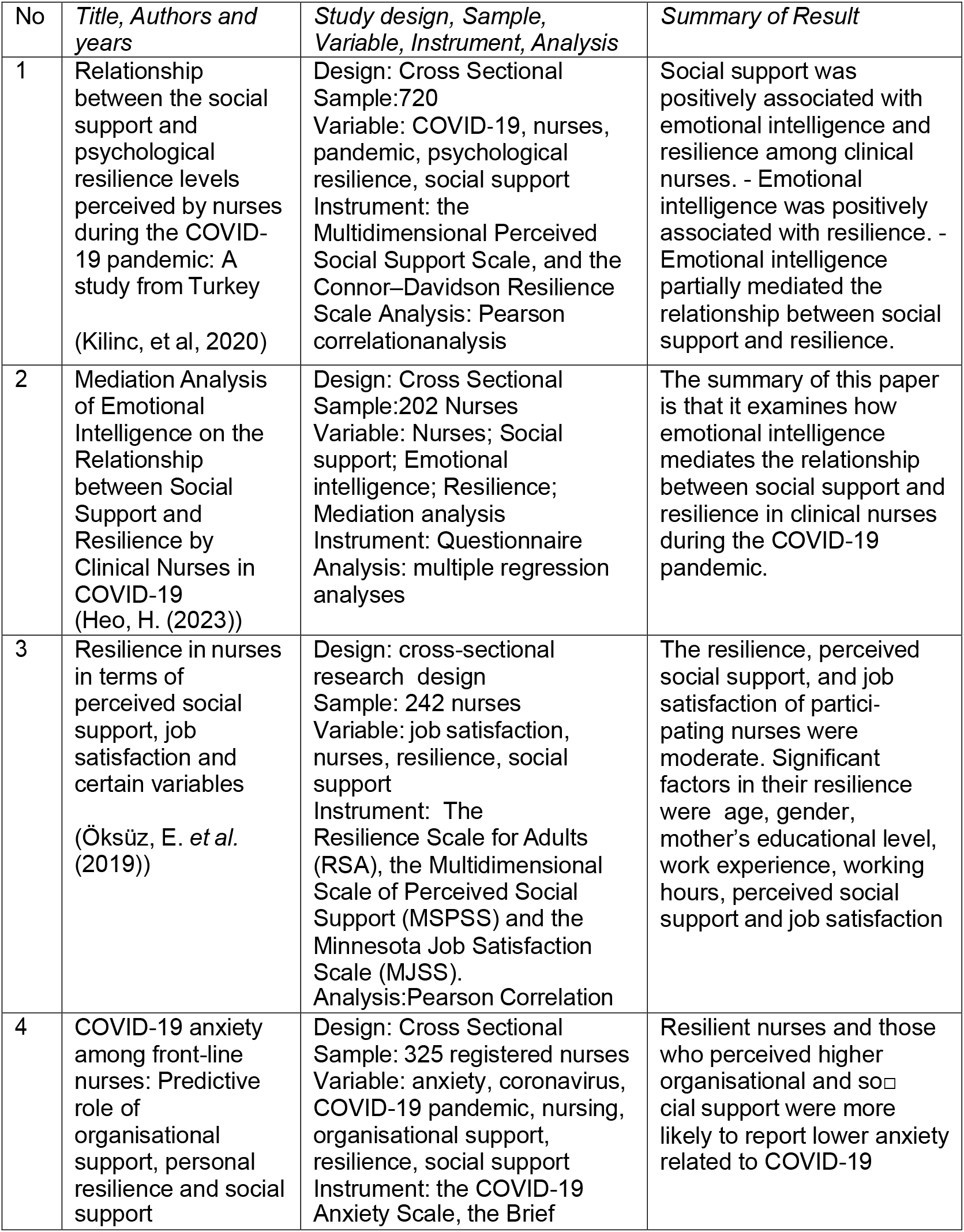
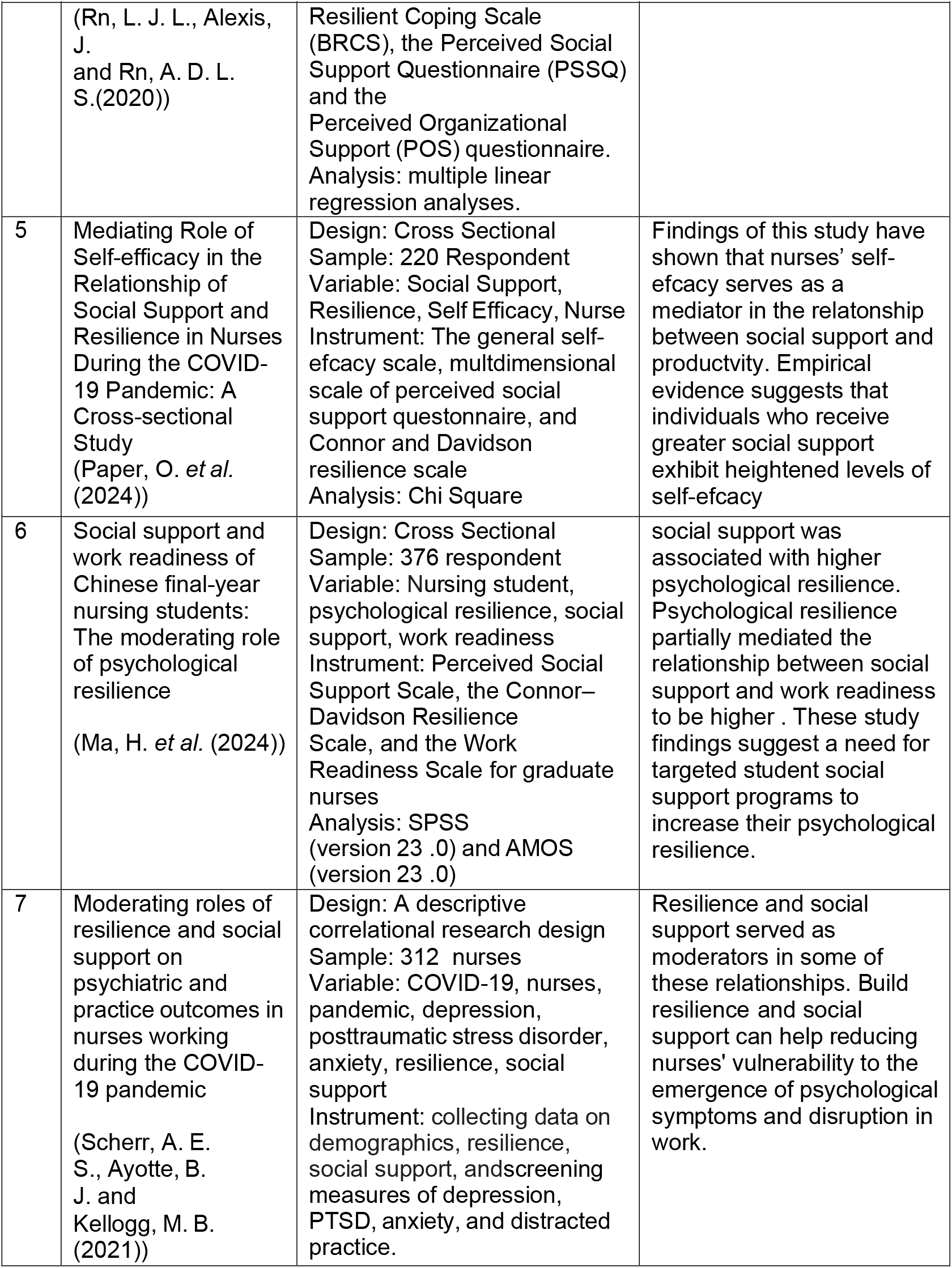
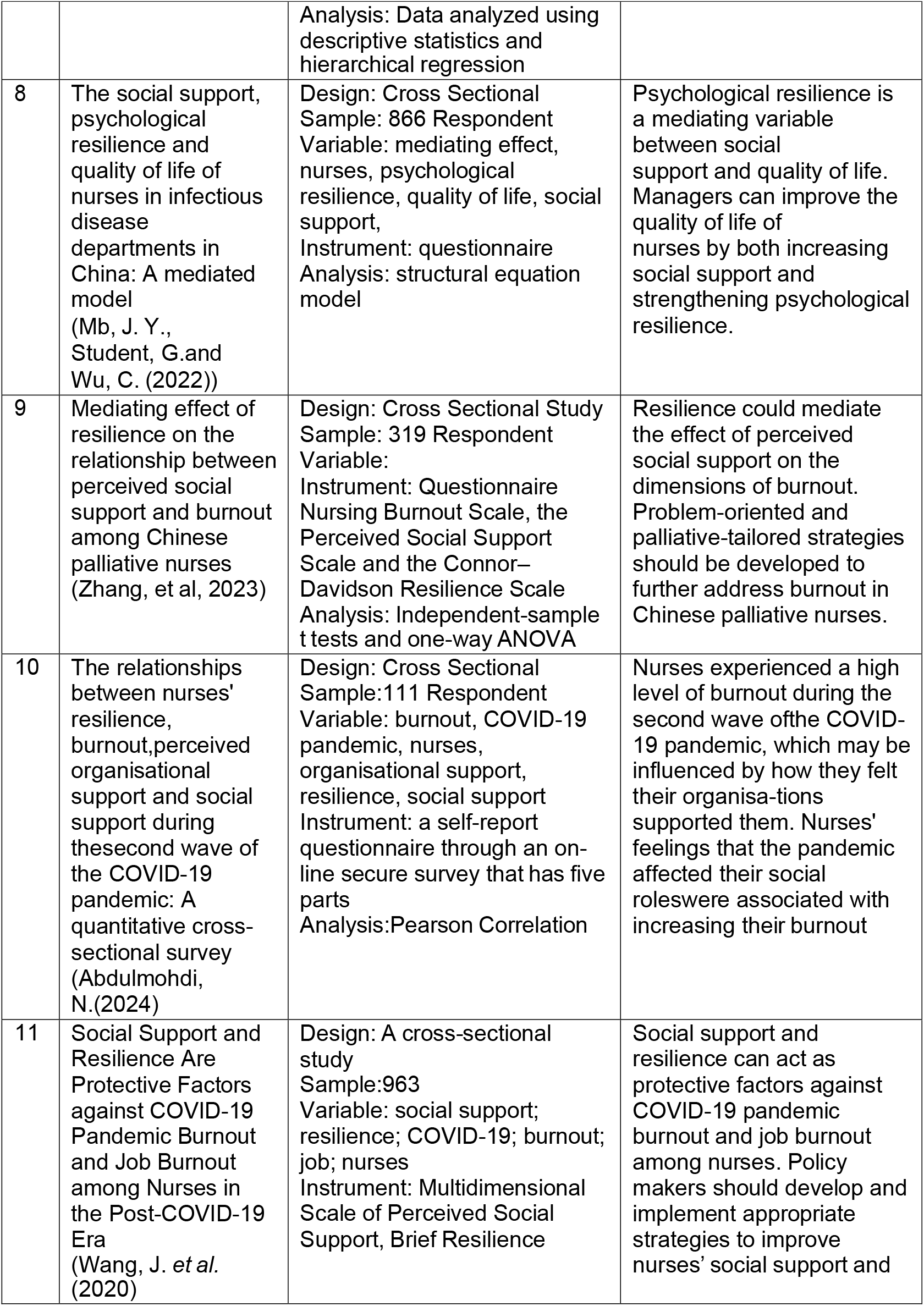
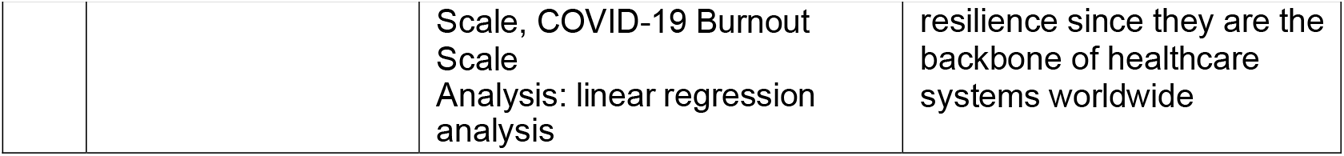
Results of Article Analysis.

| No | *Title, Authors and years* | *Study design, Sample, Variable, Instrument, Analysis* | *Summary of Result* |
| --- | --- | --- | --- |
| 1 | Relationship between the social support and psychological resilience levels perceived by nurses during the COVID-19 pandemic: A study from Turkey<br>(Kilinc, et al, 2020) | Design: Cross Sectional<br>Sample:720<br>Variable: COVID-19, nurses, pandemic, psychological resilience, social support<br>Instrument: the Multidimensional Perceived Social Support Scale, and the Connor–Davidson Resilience Scale<br>Analysis: Pearson correlation analysis | Social support was positively associated with emotional intelligence and resilience among clinical nurses. - Emotional intelligence was positively associated with resilience. - Emotional intelligence partially mediated the relationship between social support and resilience. |
| 2 | Mediation Analysis of Emotional Intelligence on the Relationship between Social Support and Resilience by Clinical Nurses in COVID-19<br>(Heo, H. (2023)) | Design: Cross Sectional<br>Sample:202 Nurses<br>Variable: Nurses; Social support; Emotional intelligence; Resilience;<br>Mediation analysis<br>Instrument: Questionnaire<br>Analysis: multiple regression analyses | The summary of this paper is that it examines how emotional intelligence mediates the relationship between social support and resilience in clinical nurses during the COVID-19 pandemic. |
| 3 | Resilience in nurses in terms of perceived social support, job satisfaction and certain variables<br>(Öksüz, E. *et al.* (2019)) | Design: cross-sectional research design<br>Sample: 242 nurses<br>Variable: job satisfaction, nurses, resilience, social support<br>Instrument: The Resilience Scale for Adults (RSA), the Multidimensional Scale of Perceived Social Support (MSPSS) and the Minnesota Job Satisfaction Scale (MJSS).<br>Analysis: Pearson Correlation | The resilience, perceived social support, and job satisfaction of participating nurses were moderate. Significant factors in their resilience were age, gender, mother's educational level, work experience, working hours, perceived social support and job satisfaction |
| 4 | COVID-19 anxiety among front-line nurses: Predictive role of organisational support, personal resilience and social support | Design: Cross Sectional<br>Sample: 325 registered nurses<br>Variable: anxiety, coronavirus, COVID-19 pandemic, nursing, organisational support, resilience, social support<br>Instrument: the COVID-19 Anxiety Scale, the Brief | Resilient nurses and those who perceived higher organisational and social support were more likely to report lower anxiety related to COVID-19 |

| No | Title, Authors and years | Study design, Sample, Variable, Instrument, Analysis | Summary of Result |
| --- | --- | --- | --- |
|  | (Rn, L. J. L., Alexis, J. and Rn, A. D. L. S.(2020)) | Resilient Coping Scale (BRCS), the Perceived Social Support Questionnaire (PSSQ) and the Perceived Organizational Support (POS) questionnaire. Analysis: multiple linear regression analyses. |  |
| 5 | Mediating Role of Self-efficacy in the Relationship of Social Support and Resilience in Nurses During the COVID-19 Pandemic: A Cross-sectional Study (Paper, O. *et al.* (2024)) | Design: Cross Sectional<br>Sample: 220 Respondent<br>Variable: Social Support, Resilience, Self Efficacy, Nurse<br>Instrument: The general self-efficacy scale, multidimensional scale of perceived social support questionnaire, and Connor and Davidson resilience scale<br>Analysis: Chi Square | Findings of this study have shown that nurses' self-efficacy serves as a mediator in the relationship between social support and productivity. Empirical evidence suggests that individuals who receive greater social support exhibit heightened levels of self-efficacy |
| 6 | Social support and work readiness of Chinese final-year nursing students: The moderating role of psychological resilience (Ma, H. *et al.* (2024)) | Design: Cross Sectional<br>Sample: 376 respondent<br>Variable: Nursing student, psychological resilience, social support, work readiness<br>Instrument: Perceived Social Support Scale, the Connor–Davidson Resilience Scale, and the Work Readiness Scale for graduate nurses<br>Analysis: SPSS (version 23 .0) and AMOS (version 23 .0) | social support was associated with higher psychological resilience. Psychological resilience partially mediated the relationship between social support and work readiness to be higher . These study findings suggest a need for targeted student social support programs to increase their psychological resilience. |
| 7 | Moderating roles of resilience and social support on psychiatric and practice outcomes in nurses working during the COVID-19 pandemic (Scherr, A. E. S., Ayotte, B. J. and Kellogg, M. B. (2021)) | Design: A descriptive correlational research design<br>Sample: 312 nurses<br>Variable: COVID-19, nurses, pandemic, depression, posttraumatic stress disorder, anxiety, resilience, social support<br>Instrument: collecting data on demographics, resilience, social support, and screening measures of depression, PTSD, anxiety, and distracted practice. | Resilience and social support served as moderators in some of these relationships. Build resilience and social support can help reducing nurses' vulnerability to the emergence of psychological symptoms and disruption in work. |
|  |  | Analysis: Data analyzed using descriptive statistics and hierarchical regression |  |
| 8 | The social support, psychological resilience and quality of life of nurses in infectious disease departments in China: A mediated model (Mb, J. Y., Student, G. and Wu, C. (2022)) | Design: Cross Sectional<br>Sample: 866 Respondent<br>Variable: mediating effect, nurses, psychological resilience, quality of life, social support,<br>Instrument: questionnaire<br>Analysis: structural equation model | Psychological resilience is a mediating variable between social support and quality of life. Managers can improve the quality of life of nurses by both increasing social support and strengthening psychological resilience. |
| 9 | Mediating effect of resilience on the relationship between perceived social support and burnout among Chinese palliative nurses (Zhang, et al, 2023) | Design: Cross Sectional Study<br>Sample: 319 Respondent<br>Variable:<br>Instrument: Questionnaire<br>Nursing Burnout Scale, the Perceived Social Support Scale and the Connor–Davidson Resilience Scale<br>Analysis: Independent-sample t tests and one-way ANOVA | Resilience could mediate the effect of perceived social support on the dimensions of burnout. Problem-oriented and palliative-tailored strategies should be developed to further address burnout in Chinese palliative nurses. |
| 10 | The relationships between nurses' resilience, burnout, perceived organisational support and social support during the second wave of the COVID-19 pandemic: A quantitative cross-sectional survey (Abdulmohdi, N.(2024) | Design: Cross Sectional<br>Sample: 111 Respondent<br>Variable: burnout, COVID-19 pandemic, nurses, organisational support, resilience, social support<br>Instrument: a self-report questionnaire through an on-line secure survey that has five parts<br>Analysis: Pearson Correlation | Nurses experienced a high level of burnout during the second wave of the COVID-19 pandemic, which may be influenced by how they felt their organisations supported them. Nurses' feelings that the pandemic affected their social roles were associated with increasing their burnout |
| 11 | Social Support and Resilience Are Protective Factors against COVID-19 Pandemic Burnout and Job Burnout among Nurses in the Post-COVID-19 Era (Wang, J. et al. (2020) | Design: A cross-sectional study<br>Sample: 963<br>Variable: social support; resilience; COVID-19; burnout; job; nurses<br>Instrument: Multidimensional Scale of Perceived Social Support, Brief Resilience | Social support and resilience can act as protective factors against COVID-19 pandemic burnout and job burnout among nurses. Policy makers should develop and implement appropriate strategies to improve nurses' social support and |
|  |  | Scale, COVID-19 Burnout Scale<br>Analysis: linear regression analysis | resilience since they are the backbone of healthcare systems worldwide |

## Results and Discussion

The results of a review of 11 articles describing relationship between social support and resilience in nurses,namely research Kilinc, et al, 2020; Heo, H. (2023); Öksüz, E. *et al*. (2019); Rn, L. J. L., Alexis, J. and Rn, A. D. L. S. (2020); Paper, O. *et al*. (2024); Ma, H. *et al*. (2024); Scherr, A. E. S., Ayotte, B. J. and Kellogg, M. B. (2021); Mb, J. Y., Student, G. and Wu, C. (2022); Zhang, et al, 2023; Abdulmohdi, N. (2024); Wang, J. *et al*. (2020).

### Ontological, Axiological, and Epistemological study of social support and resilience in nurses

**The ontological study** of social support and resilience, social support plays a significant role in enhancing nurse resilience, which is crucial for coping with the demanding nature of their profession. The relationship between social support and resilience is multifaceted and involves various mediating factors.

Direct Impact of Social Support: Social support has a direct positive effect on nurse resilience. Nurses who receive higher levels of social support tend to exhibit greater resilience, which helps them manage stress and maintain their well-being (Heo, H. 2023; Kilinc, et al, 2020; Paper, O. *et al*. (2024). Peer support, in particular, is highlighted as a significant factor in helping nurses with challenging situations, such as dealing with patient deaths andproviding hospice care (Moisoglou, I. *et al*. (2024).

**The Axiological study** of social support and resilience, Self-Efficacy: Self-efficacy mediates the relationship between social support and resilience. Higher social support boosts self-efficacy, which in turn enhances resilience (Heo, H. 2023)

.Emotional Intelligence: Emotional intelligence partially mediates the relationship between social support and resilience, suggesting that emotionally intelligent nurses can better leverage social support to build resilience (Kilinc, et al, 2020). Psychological Resilience: Psychological resilience itself acts as a mediator between social support and various outcomes, such as work readiness and quality of life (Wang, J. *et al*. (2020)).

Increased social support and resilience are associated with better job performance and reduced burnout among nurses. This is particularly important during crises like the COVID-19 pandemic, where nurses face heightened stress and burnout (Scherr, A. E. S., Ayotte, B. J. and Kellogg, M. B. (2021). Social support and resilience can act as protective factors against job burnout, highlighting the need for supportive work environments (Scherr, A. E. S., Ayotte, B. J. and Kellogg, M. B. (2021).

Promoting Social Support: Nursing administrators should foster a supportive work environment by encouraging peer support and mentorship programs (Abdulmohdi, N. (2024; Kilinc, et al, 2020). This can help build a strong support network among nurses, enhancing their resilience. Implementing organizational interventions that increase social support and provide psychological and mental health services can help reduce anxiety and improve overall well-being (Scherr,et al. (2021).

Training and Development: Training programs focused on building self-efficacy and emotional intelligence can further enhance the positive effects of social support on resilience (Abdulmohdi, N. (2024); Heo, H. 2023). Resilience training programs and strategies tailored to the specific needs of nurses, such as those working in palliative care or during pandemics, can be particularly beneficial (Huang, F. *et al*.*2024)*. In summary, social support significantly influences nurse resilience through direct effects and various mediating factors. Enhancing social support within healthcare settings can lead to improved resilience, better job performance, and reduced burnout among nurses. Based on the user’s query, the impact of social supporton nurse resilience in thecontext of workplace stress can be explored using insights fromthe provided abstracts.

**The Epistemiological study** of social support and resilience, Social support has a significant positive impact on nurse resilience, as evidenced by multiple studies (Abdulmohdi, N. (2024); Heo, H. 2023; Kilinc, et al, 2020). The relationship between social support and nurse resilience is particularly crucial during stressfulsituations, such as the COVID-19 pandemic (Abdulmohdi, N. (2024); Heo, H. 2023; Kilinc, et al, 2020). The presence of social support directly contributes to increased resilience among nurses, highlighting the importance of this factor in mitigating workplace stress Abdulmohdi, N. (2024); Heo, H. 2023; Kilinc, et al, 2020;Choi, B, 2018)

The key components of social support that contribute to nurse resilience include coworker support, friend support, and organizational support (Abdulmohdi, N. (2024); Heo, H. 2023; Kilinc, et al, 2020; Moisoglou, I. *et al*. (2024).Coworker support has been found to have a significant impact on nurse resilience, with its effect being fully mediated by self-efficacy (Moisoglou, I. *et al*. (2024). Friend support has a direct positive effect on self-efficacy and an indirect effect on nurse resilience, emphasizing the multifaceted nature of social support in enhancing resilience (Moisoglou, I. *et al*. (2024)). Organizational support also plays a crucial role in bolstering nurse resilience, especially in the context of the COVID-19 pandemic (Abdulmohdi, N. (2024); Heo, H. 2023; Kilinc, et al, 2020).

Organizational policies and practices can enhance social support for nurses by fostering a positive work climate, promoting effective mentorship programs, and providing resources to protect the well-being of nursing staff (Abdulmohdi, N. (2024); Heo, H. 2023; Kilinc, et al, 2020). Nurse managers should consider measures to protect their staff, such as developing effective mentorship programs and promoting a positive work climate (Abdulmohdi, N. (2024).It is essential for nursing managers to determine the social support resources of nurses, especially during epidemic periods,and to conduct individual and institutional studies to increase nurses’ psychologicalresilience (Wang, J. *et al*. (2020).

## Conclusion

The perception of social support and its impact on resilience may vary between novice and experienced nurses, but this specific comparison is not directly addressed in the provided abstracts. In conclusion, the relationship between social support and nurse resilience is well-supported by the provided abstracts, highlighting the significant positive impact of social support on nurse resilience, the key components of social support, and the potential for organizational policies and practices to enhance social support for nurses. However, the specific differences in the perception of social support and its impact on resilience between novice and experienced nurses.

## Data Availability

All data produced in the present work are contained in the manuscript

## REFERENCES

Abdulmohdi, N. (2024) ‘The relationships between nurses’ resilience, burnout, perceivedorganisational support and social support during the second wave of the COVID-19 pandemic : A quantitative cross-sectional survey’, (October 2023), pp. 1–12. doi: 10.1002/nop2.2036.

Almazan, J. U. et al. (2019) ‘International Journal of Disaster Risk Reduction Disaster-related resiliency theory among older adults who survived Typhoon Haiyan’, InternationalJournal of Disaster Risk Reduction, 35(February), p. 101070. doi: 10.1016/j.ijdrr.2019.101070.

Cao, J. et al. (2024) ‘The association between perceive social support and post-traumatic stress disorder symptoms among medical staff in Hubei, China : a chain mediating effectof resilience and positive coping’.

Chigwedere, O. C. et al. (2021) ‘The Impact of Epidemics and Pandemics on the Mental Health of Healthcare Workers: A Systematic Review’.

Doctor, W. Z., Doctor, R. M. and Doctor, X. S. (2020) ‘Burnout in nurses working in China : A national questionnaire survey’, (September), pp. 1–9. doi: 10.1111/ijn.12908.

Heo, H. (2023) ‘COVID-19 상황의 임상간호사가 지각하는 사회적 지지와 회복탄력성의 관계에서 감성지능의 매개효과 Mediation Analysis of Emotional Intelligence on the Relationship between Social Support and Resilience by Clinical Nursesin COVID-19’, 29(3), pp. 181–190.

Huang, F. et al. 2024. ‘Acknowledgements We gratefully acknowledge all the study participants, without them, it is not possible to complete these projects.’, pp. 1–21. doi: 10.1111/nhs.12859.

Ma, H. et al. (2024) ‘Social support and work readiness of Chinese final-year nursing students : The moderating role of psychological resilience’. doi: 10.1080/14330237.2023.2291231.

Mb, J. Y., Student, G. and Wu, C. (2022) ‘The social support, psychological resilience andquality of life of nurses in infectious disease departments in China : A mediated model’, (September), pp. 4503–4513. doi: 10.1111/jonm.13889.

‘Mediating effect of resilience on the relationship between perceived social support and burnout among Chinese palliative nurses’ (2024), p. 16532. doi: 10.1111/jocn.16532.

Moisoglou, I. et al. (2024) ‘Social Support and Resilience Are Protective Factors against COVID-19 Pandemic Burnout and Job Burnout among Nurses in the Post-COVID-19 Era’.

Nurs, M. H., Terms, W. and Versions, S. (2020) ‘Nurse Resilience: A Concept Analysis’, pp.553–575.

Öksüz, E. et al. (2019) ‘Resilience in nurses in terms of perceived social support, job satisfaction and certain variables’, Journal of Nursing Management, 27(2), pp. 423–432. doi: 10.1111/jonm.12703.

Paper, O. et al. (2024) ‘Mediating Role of Self-efficacy in the Relationship of Social Support and Resilience in Nurses During the COVID-19 Pandemic: A Cross-sectional Study’, 34(3), pp. 229–236.

Rn, L. J. L., Alexis, J. and Rn, A. D. L. S. (2020) ‘COVID-19 anxiety among front-line nurses : Predictive role of organisational support, personal resilience and social support’, (August), pp. 1653–1661. doi: 10.1111/jonm.13121.

Scherr, A. E. S., Ayotte, B. J. and Kellogg, M. B. (2021) ‘Moderating Roles of Resilience and Social Support on Psychiatric and Practice Outcomes in Nurses Working During the COVID-19 Pandemic’. doi: 10.1177/23779608211024213.

Wang, J. et al. (2020) ‘International Journal of Nursing Studies Factors associated with compassion satisfaction, burnout, and secondary traumatic stress among Chinese nurses in tertiary hospitals : A cross-sectional study’, 102, pp. 1–8. doi: 10.1016/j.ijnurstu.2019.103472.

